# Spread of COVID-19 in India: A Simple Algebraic Study

**DOI:** 10.1101/2020.05.10.20097691

**Authors:** Sudipto Roy

**Affiliations:** Department of Physics, St. Xavier’s College, Kolkata, West Bengal, India

**Keywords:** COVID-19, SARS-CoV-2, Pandemic, India, Social Distancing, Epidemiology, Mathematical Model.

## Abstract

The number of patients, infected with COVID-19, began to increase very rapidly in India from March 2020. The country was put under lockdown from 25 March 2020. The present study is aimed at providing a simple algebraic analysis of the trend that is evident in the spread of the disease in this part of the world. The purpose of this algebraic approach is to simplify the calculation sufficiently by deviating from the standard techniques that are conventionally used to construct mathematical models of epidemics. The predictions, obtained from this algebraic study, are found to be in reasonable agreement with the recorded data. Using this mathematical formulation we have determined the time variation of the number of asymptomatic patients, who are believed to play a major role in spreading the disease. We have discussed the effect of lockdown in reducing the rate of transmission of the disease. On the basis of the proposed models, predictions have been made regarding the possible trend of the rise in the number of cases beyond the withdrawal of lockdown. All these things have been calculated by using very simple mathematical expressions which can be easily understood and used by those who have a rudimentary knowledge of algebra.

## 1. Introduction

In early December 2019, a cluster of cases of pneumonia had been reported in Wuhan, Hubei province of China. A few days later, the health authorities of that country declared that this cluster was associated with a newly discovered coronavirus (SARS-CoV-2) and the infectious disease caused by it was named coronavirus disease 2019 (COVID-19) by the World Health Organization (WHO) [1, 2]. This outbreak of novel coronavirus pneumonia has been declared a public health emergency of international concern by WHO. As of 6:37 pm CEST, 10 May 2020, there have been 3,925,815 confirmed cases of COVID-19, including 274,488 deaths, reported to WHO globally [3]. According to WHO, no pharmaceutical products have yet been shown to be safe and effective for the treatment of COVID-19. In-vitro studies have shown that chloroquine, an immune-modulant drug, which has been traditionally used for the treatment of malaria, is effective in reducing viral replication in infections including the SARS-associated coronavirus (CoV) and MERS-CoV [4, 5]. It has been revealed by the current studies that respiratory symptoms of COVID-19 such as fever, dry cough and dyspnea are the most common manifestations, quite similar to severe acute respiratory syndrome (SARS) in 2003 and Middle East respiratory syndrome in 2012 (MERS), which firmly indicates the droplet transmission and contact transmission of the virus. Apart from the typical respiratory disorder, there are less common features like diarrhea, nausea, vomiting, and abdominal discomfort that have been found significantly in different degrees among different study populations [6]. Recent evidences have revealed that COVID-19 virus is transmitted between people through respiratory droplets and contact routes [7-11]. Transmission through droplets takes place when a person is in close contact (within 1 metre) with another person, who has developed respiratory symptoms (coughing or sneezing) due to COVID-19 infection, and is thereby at risk of having his/her mucosae (mouth and nose) or conjunctiva (eyes) exposed to potentially infective respiratory droplets (which are generally known to be greater than 5-10 μm in diameter). Droplet transmission can also occur through fomites in the immediate vicinity around the infected person [12]. Thus, the transmission of the virus that causes COVID-19 can occur by direct contact with an infected person and indirect contact with surfaces in his/her immediate environment or with the objects that have been used on the infected person (e.g. stethoscope or thermometer). Airborne transmission of the virus is not the same as droplet transmission. It is caused by the microbes within droplet nuclei, which are generally like particles less than 5μm in diameter, and which are released by the evaporation of larger droplets or exist within dust particles. These particles may remain in the air for long periods of time and they can be transmitted to persons over distances greater than 1 metre [13].

The first case of COVID-19 infection was reported in India on 30 January 2020. As of 8:00 am IST, 10 May 2020, a total of 62,939 cumulative cases of infection including 41,472 active cases, 19,357 recoveries and 2,109 deaths in the country have been confirmed by the Ministry of Health and Family Welfare [14]. So far, the government has issued necessary guidelines and taken several measures to spread awareness regarding COVID-19 and also to enforce social distancing of its citizens to break the chain of transmission of the disease. On 24 March 2020, a nationwide lockdown was announced for a period from 25 March 2020 to 14 April 2020. On 14 April 2020, this lockdown was extended till 03 May 2020. On 01 May 2020 it was further extended till 17 May 2020.

Chatterjee et al. have carried out a detailed study to accumulate evidence that can guide research activities towards the prevention and control such a pandemic spreading so rapidly in India [15]. In another study, Agarwal et al have elaborately discussed the necessity for a proper medical infrastructure to be built up in India to tackle the flow of patients and to ensure the safety of the healthcare workers [16]. Through a detailed mathematical analysis, Mandal et al have discussed the policies to be implemented to prevent the spread of the virus by community transmission [17]. Some other mathematical models, based on several standard theories, have been constructed to predict the number of infections of COVID-19 in India with sufficient accuracy [18-25]. These models are expected to serve as efficient tools which would certainly help the policy makers of the country, at different levels, to make proper plans to prevent the spread of this disease.

In a previously published article, based on a mathematical model, we showed the effect of imposition of lockdown in reducing the rate of rise in the number of infections [26]. It was a dynamical model which was based on a differential equation that was formulated to find the time variation of the number of asymptomatic patients, from which, the number of symptomatic cases was estimated. In the present study, we have constructed the entire mathematical structure upon a simple algebraic equation. The purpose of choosing this method is to make the article comprehensible to the policy makers of the country who come from various educational backgrounds. The models that are based on calculus lead to very accurate results or predictions but they are sometimes very difficult to understand for those who are not sufficiently trained in mathematics. In most of the cases, the conventional models do not lead to mathematical expressions from which predictions can be made. One needs to do numerical calculations (rather than analytical) to arrive at a prediction. In view of the severity and urgency of the crisis caused by COVID-19 outbreak, mathematical models should be created in a simple way so that the can be easily understood, amended or modified if necessary and applied extensively by the persons who are responsible for decision making regarding infrastructural arrangements and also the formulation and implementation of rules to be imposed upon the society to ensure social distancing. The calculations involved in the present study are extremely simple in comparison to the ones where one needs to solve a set of coupled differential equations keeping under consideration various factors connected to the society and the constraints of the actual situations caused by the pandemic and the measures to control it.

In the present article, we have discussed the step by step construction of an algebraic structure that allows one to derive an expression representing the time evolution of the number of asymptomatic patients in the country. Like our previously published article, which was based on calculus, this model has an underlying assumption that, due to the lack of tests in sufficient numbers, the statistics regarding the number of patients infected, as declared by the government, are actually about the number of symptomatic patients. After detection, most of them are put under isolation, during which they are not much capable of spreading the disease to other persons. Therefore, the asymptomatic carriers can be regarded as the main agents of transmission of the virus in the society. The present study is based on three models whose unknown parameters have been determined by fitting these models to the actual data of COVID-19 infections in India. For this purpose we have used the statistics of the number of infected persons in India, during the period from 01 March 2020 to 03 May 2020, obtained from the government sources [14]. Using these three models we have determined the time evolution of the number of infected persons over this period. We have graphically shown the positive impact of the imposition of lockdown throughout the country. The present study makes predictions regarding the number of infected cases beyond 17 May 2020, the date up to which the lockdown will continue as per the last announcement made on 01 May 2020. These models show very clearly that a high degree of social distancing has to be maintained to slow down or prevent the transmission of the disease in the country.

## 2. Mathematical Models

Let *y*_1_ and *y*_2_ be the numbers of asymptomatic patients on the first and the second days of the span of time under consideration. We propose to express *y*_2_ in terms of *y*_1_ in the following way.

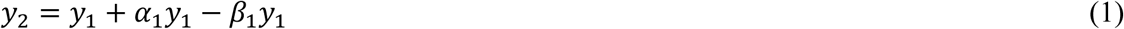

In equation (1), *α*_1_ is the average number of persons who become infected with COVID-19, after coming in close contact with each of these *y*_1_ carriers of the disease, on the first day. Here *β*_1_ denotes the fraction of *y*_1_ who have undergone a transformation from asymptomatic to symptomatic modes on the first day. It is assumed in the present model that a patient, after being identified as symptomatic, is put into absolute isolation from the society. It prevents the patient completely from playing any role in the transmission of the disease. It is also assumed that no new asymptomatic or symptomatic carrier has entered the geographical region under consideration, during the entire span of time for which this study has been conducted.

The subscript of *α*_1_ is connected to the serial number of the day concerned. When a lockdown is imposed, the social mixing pattern changes significantly, resulting in a change in the number of persons coming in contact with an asymptomatic carrier. Let us consider a combination of three consecutive periods, of *d*_1_, *d_2_* & *d*_3_ days respectively, during which we have *no-lockdown, lockdown* and again *no-lockdown* situations successively in a country. The time dependence of *α_n_* (i.e. dependence upon *n*), under such situations, can be expressed as,

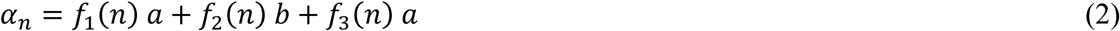

Here, the value of each of these three functions, *f*_1_(*n*)*, f*_2_(*n*) and *f*_3_(*n*), is unity over the periods of *d*_1_*, d*_2_ & *d*_3_ days respectively, and zero otherwise. Thus, the values of *α_n_* would be equal to the constants, and again, respectively, during these three periods.

We have *α_n_ = a* for 1 ≤ *n* ≤ *d*_1_, *α_n_* = *b* for *d*_1_ < *n* ≤ *d*_2_, and *α_n_* = *a* for *d*_2_ < *n* ≤ *d*_3_. Here *b* and *a* can be regarded, respectively, as the measures of social distancing during *lockdown* and *no-lockdown* periods. They decrease as social distancing increases.

Like *α*_1_, the subscript of *β*_1_ corresponds to the serial number of the day under consideration. Its time dependence is due to an assumption that the number of asymptomatic patients may increase at such a rate that, the fraction of them turning into symptomatic ones on a certain day, cannot have a constant value. One of the three forms of *β_n_*, introduced in the present paper, has been assumed to have a constant value (denoted by *γ* in section 2.3), which might be regarded as a kind of time-average of that fraction.

A generalized version of equation (1) can be expressed as,

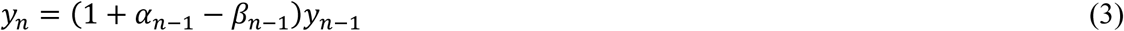

For *n =* 2, one obtains equation (1) from equation (3). Putting *n = n* − 1 in equation (3), one gets,

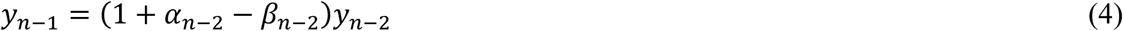

Substituting for *y_n_*_−_*_i_* in equation (3) from equation (4), one obtains,

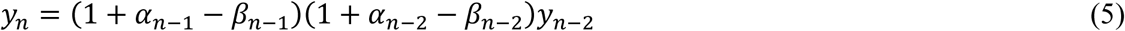

Continuing in this fashion, equation (3) can be written as,

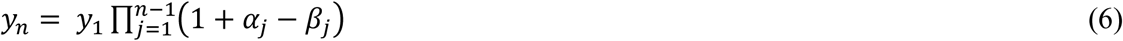

The number of cases (*x_k_*) that become symptomatic from the asymptomatic type, on the *k^th^*
day, is then given by,

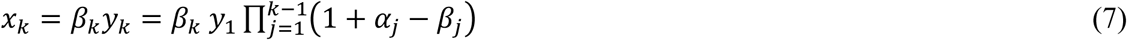

Therefore, the total number of symptomatic cases recorded till the *n^th^* day is given by,

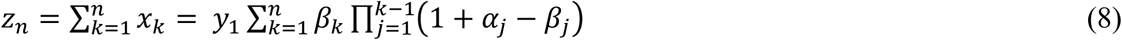

In equations (6), (7) and (8) *α_j_ = f*_1_(*j*) *a* + f_2_(*j*)*b* + *f*_3_(*j*)*a* as per equation (2). If the third
phase is absent we have *α_j_ = f*_1_(*j*) *a* + f_2_(*j*)*b*. There can be many such phases coming one
after another. For a process, with *L* number of such phases, *α_j_* is given by,

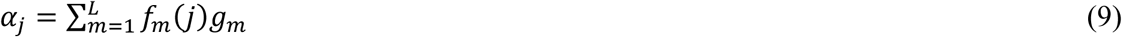

Here *g_m_ = a* for the odd values of *m* and *g_m_ = b* for the even values of *m*.

Equation (8) actually gives us the value that needs to be compared with the number that is declared by the government as the total number of confirmed COVID-19 cases registered in the country till the n^th^ day. It is the cumulative count of asymptomatic cases till that day.

Without any imposition of lockdown, over the entire period under study, we must have

*α_j_ = a*, according to equation (2). Equation (8) will then have the following form.

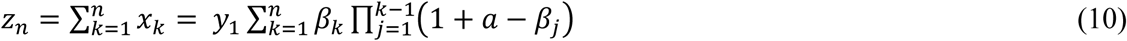

Relative proportions of the symptomatic and asymptomatic patients, denoted by *P(z_n_)* and *P(y_n_)* respectively, are given by the following two expressions.

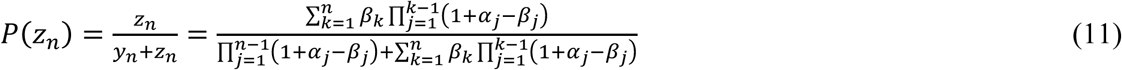

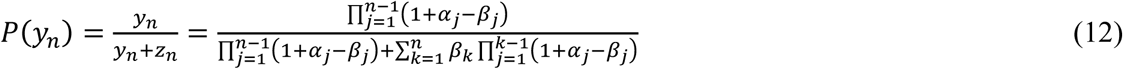

*P*(*z_n_*) and *P*(*z_y_*) can be expressed in percentages by multiplying their expressions with 100. As an average estimate one can say that, for each symptomatic case there are *y_n_/z_n_* or *P*(*y_n_*) */P*(*z_n_*) number of asymptomatic cases, which remain mostly undetected in India due to lack of testing facilities.

Combining the numbers of symptomatic and asymptomatic cases, one gets the total number of cumulative infections in the country. Dividing this value by the present population of India (*N*), one can get the fraction of the population infected with COVID-19. This fraction, denoted by *F_n_* here, is given by,

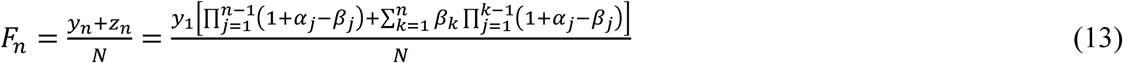

*F_n_* can be expressed in percentages by multiplying its expressions with 100. *N* = 1.360 × 10^9^.

For the present article, we have assumed three functional forms of *β_j_* described in the following three models.

### 2.1 Model-1: Exponential expression for *β_j_*

Here we assume the following relation for *β_j_*.

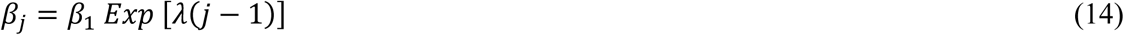

In equation (14), *β*_1_ and *λ* are constants. The parameter *λ* determines how fast *β_j_* changes for any change of *j*. Substituting this *β_j_* into equations (6), (8) and (10), respectively, we get,

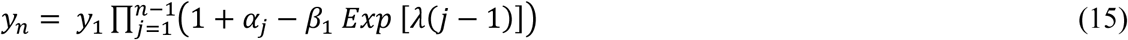

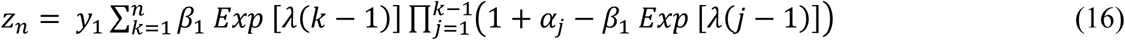

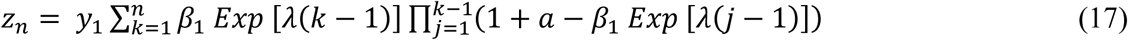

### 2.2 Model-2: Power law expression for *β_j_*

Here we assume the following relation for *β_j_*.

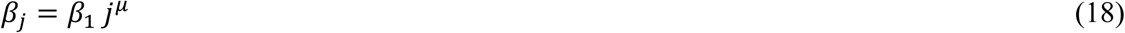

In equation (18), *β*_1_ and *μ* are constants. The parameter *μ* determines how fast *β_j_* changes for any change of *j*. Substituting this *β_j_* into equations (6), (8) and (10), respectively, we get,

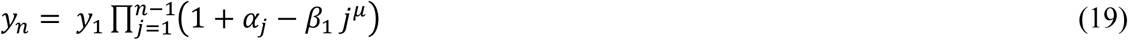

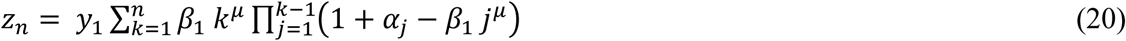

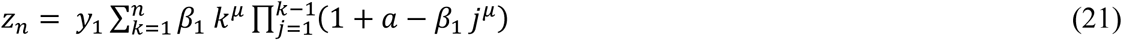

### 2.3 Model-3: A Constant *β_j_*

Here we consider the following form for *β_j_*.

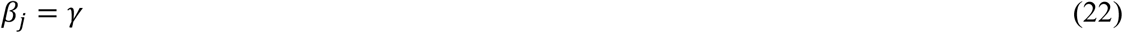

In equation (22), *γ* is a constant. Substituting this *β_j_* into equations (6), (8) and (10), respectively, we get,

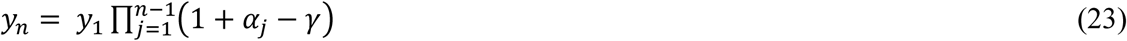

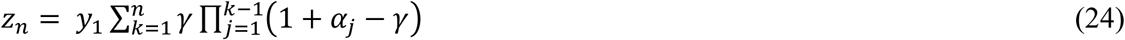

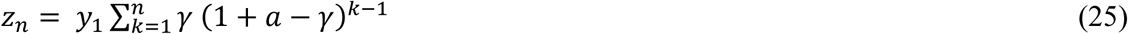

For the present study we have used the following expressions for *f*_1_(*n*), *f*_2_(*n*) and *f*_3_(*n*) respectively, which are required for equations (2) and (9).

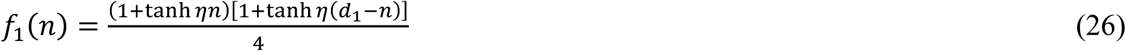

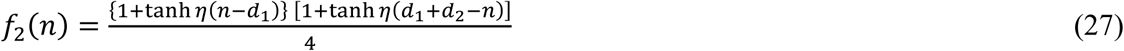

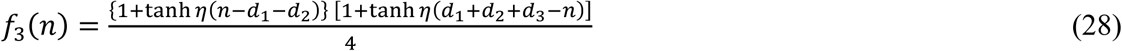

According to the definitions of *f*_1_(*n*), *f*_2_(*n*) and *f*_3_(*n*), discussed previously, they must
behave like rectangular pulses of unit heights. For this purpose, one must choose the value of the constant *η* to be sufficiently large in comparison to the values of *d*_1_, *d*_2_ and *d*_3_.

## 3. Graphical Interpretation

Figure 1 shows the time variation of the number of symptomatic patients (*z_n_*). The black circles represent the data regarding the confirmed COVID-19 cases registered in India from 01 March 2020 (i.e. *n* = 1) to 03 May 2020 (i.e. *n* = 64), as obtained from the government sources [14]. The red circles represent the values prdicted by Model-1 of the present study. The predictions from the model are in reasonable agreement with the recorded data for a certain set of parameter values which are, *y*_1_ = 100, *a* = 0.2, *b* = 0.125, *β*_1_ = 0.022 and *λ =* −0.008. These values have been used in the present study for Model-1.

**Figure 1:**
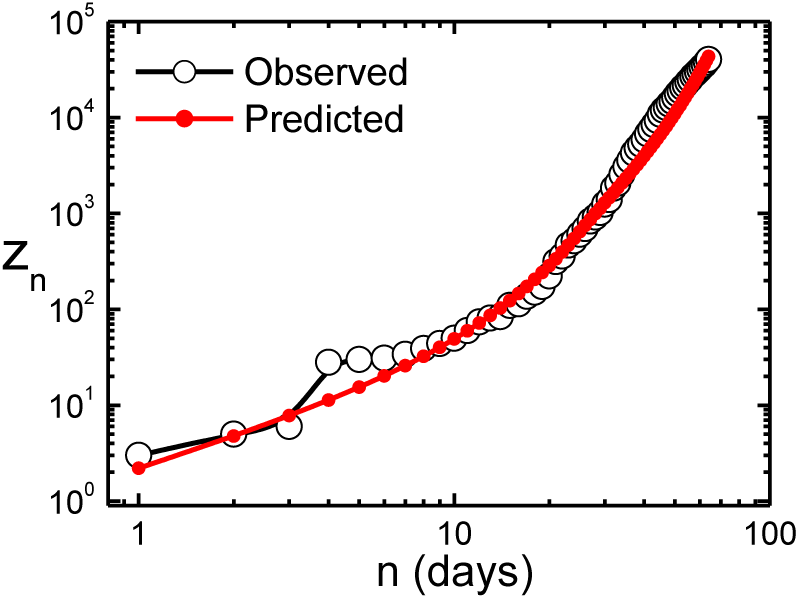
Variation of the number of symptomatic patients as a function of time. The black circles represent the numbers of confirmed COVID-19 cases registered in India from 01 March 2020 (i.e. *n* = 1) to 03 May 2020 (i.e. *n* = 64). The red circles represent the values predicted by Model-1 of the present study.

Figure 2 shows the time variation of the number of symptomatic patients (*z_n_*). The black circles represent the data regarding the confirmed COVID-19 cases registered in India from 01 March 2020 (i.e. *n* = 1) to 03 May 2020 (i.e. *n* = 64), as obtained from the government sources [14]. The red circles represent the values predicted by Model-2 of the present study. The predictions from the model are in reasonable agreement with the recorded data for a certain set of parameter values which are, *y*_1_ = 100, *a* = 0.2, *b* = 0.118, *β*_1_ = 0.03 and *μ =* − 0.1. These values have been used in the present study for Model-2.

**Figure 2:**
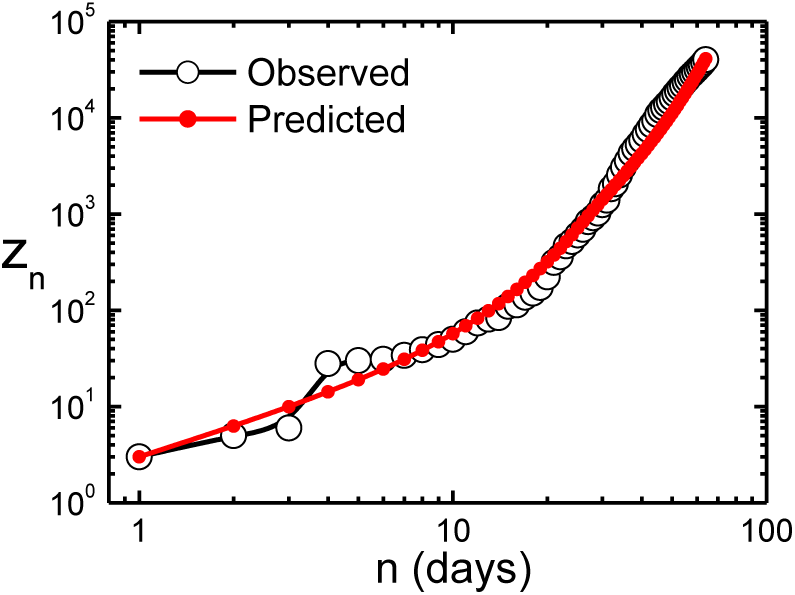
Variation of the number of symptomatic patients as a function of time. The black circles represent the numbers of confirmed COVID-19 cases registered in India from 01 March 2020 (i.e. *n* = 1) to 03 May 2020 (i.e. *n* = 64). The red circles represent the values predicted by Model-2 of the present study.

Figure 3 shows the time variation of the number of symptomatic patients (*z_n_*). The black circles represent the data regarding the confirmed COVID-19 cases registered in India from 01 March 2020 (i.e. *n* = 1) to 03 May 2020 (i.e. *n* = 64), as obtained from the government sources [14]. The red circles represent the values predicted by Model-3 of the present study. The predictions from the model are in reasonable agreement with the recorded data for a certain set of parameter values which are, *y*_1_ = 100, *a* = 0.2, *b* = 0.116, *γ*= 0.03. These values have been used in the present study for Model-3.

**Figure 3:**
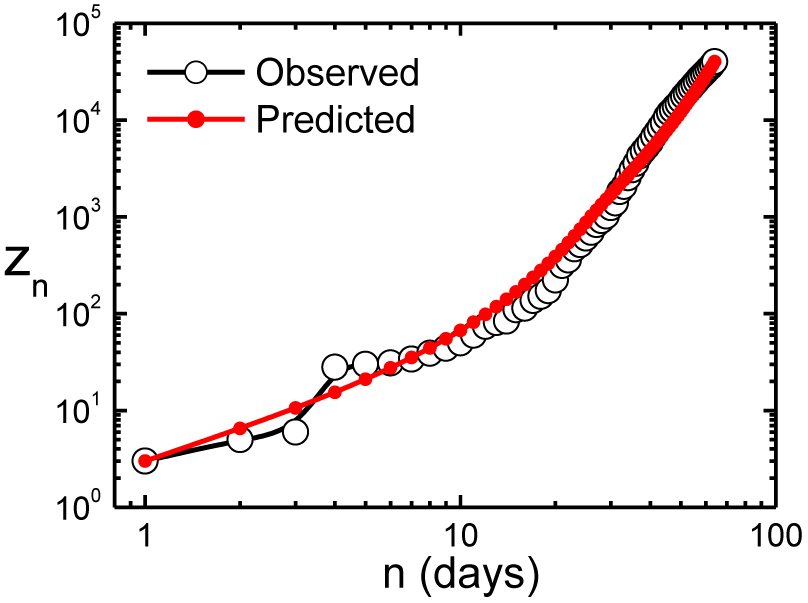
Variation of the number of symptomatic patients as a function of time. The black circles represent the numbers of confirmed COVID-19 cases registered in India from 01 March 2020 (i.e. *n* = 1) to 03 May 2020 (i.e. *n* = 64). The red circles represent the values predicted by Model-3 of the present study.

Figure 4 depicts the time evolution of the number of asymptomatic and symptomatic cases, over the period from 01 March 2020 (i.e. *n* = 1) to 03 May 2020 (i.e. *n* = 64), based on Model-1, using the parameter values obtained by fitting this model to the actual data (Fig. 1). Lockdown was in effect from 25 March 2020. The impact of lockdown is evident from the reduction of slope of both curves.

**Figure 4:**
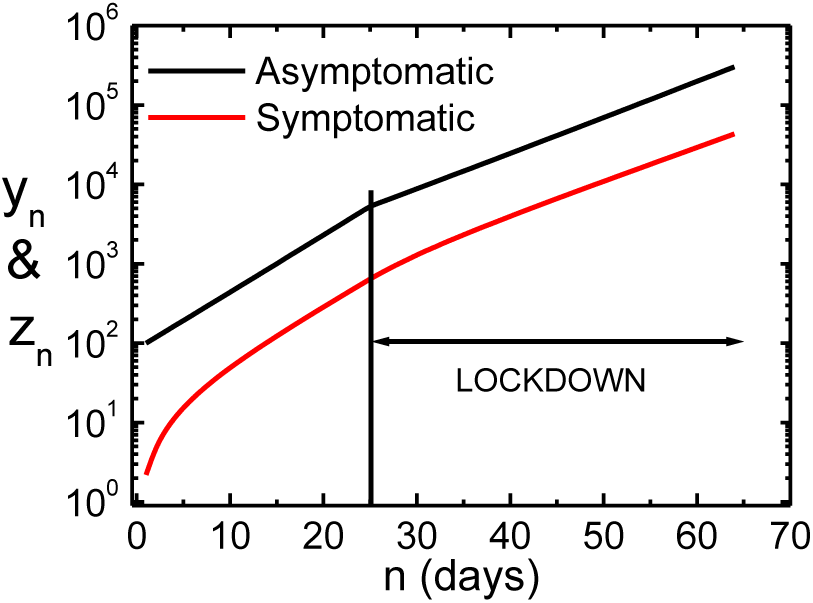
Variation of the numbers of asymptomatic (black) and symptomatic (red) patients, denoted by *y_n_* and *z_n_* respectively, as functions of time, over the period from 01 March 2020 (i.e. *n* = 1) to 03 May 2020 (i.e. *n* = 64), from Model-1, using the parameter values obtained by fitting the model to the actual data (Fig. 1).

Figure 5 shows the time variation of the number of asymptomatic and symptomatic cases, over the period from 01 March 2020 (i.e. *n* = 1) to 03 May 2020 (i.e. *n* = 64), based on Model-2, using the parameter values obtained by fitting this model to the actual data (Fig. 2). Lockdown was in effect from 25 March 2020. The impact of lockdown is evident from the reduction of slope of both curves.

**Figure 5:**
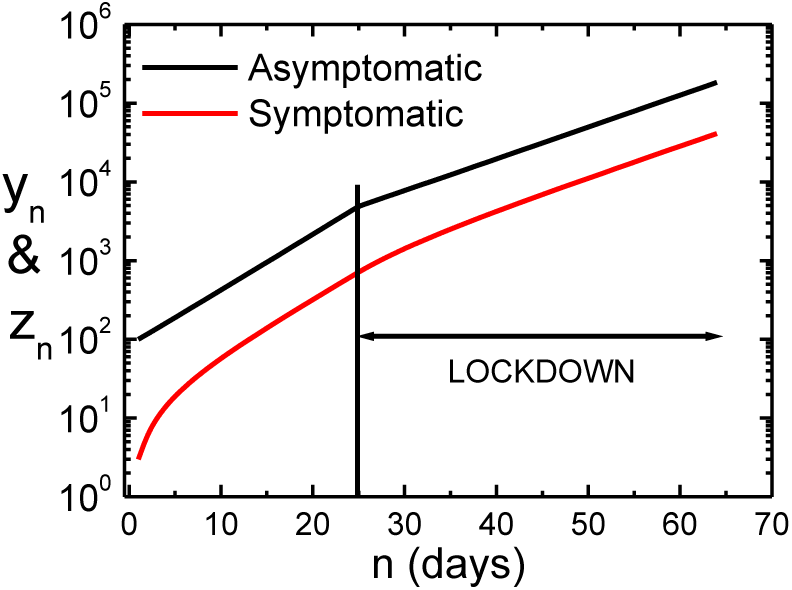
Variation of the numbers of asymptomatic (black) and symptomatic (red) patients, denoted by *y_n_* and *z_n_* respectively, as functions of time, over the period from 01 March 2020 (i.e. *n* = 1) to 03 May 2020 (i.e. *n* = 64), from Model-2, using the parameter values obtained by fitting the model to the actual data (Fig. 2).

Figure 6 depicts the time evolution of the number of asymptomatic and symptomatic cases, over the period from 01 March 2020 (i.e. *n* = 1) to 03 May 2020 (i.e. *n* = 64), based on Model-3, using the parameter values obtained by fitting this model to the actual data (Fig. 3). Lockdown was in effect from 25 March 2020. The impact of lockdown is evident from the reduction of slope of both curves.

**Figure 6:**
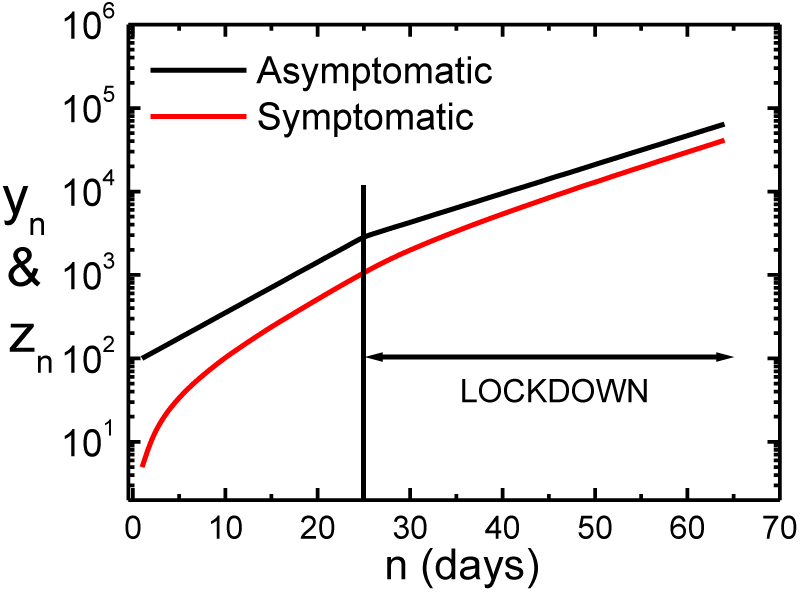
Variation of the numbers of asymptomatic (black) and symptomatic (red) patients, denoted by *y_n_* and *z_n_* respectively, as functions of time, over the period from 01 March 2020 (i.e. *n* = 1) to 03 May 2020 (i.e. *n* = 64), from Model-3, using the parameter values obtained by fitting the model to the actual data (Fig. 3).

Figure 7 shows the variation of the number of symptomatic patients as a function of time, with and without the imposition of lockdown, over the period from 01 March 2020 (i.e. *n* = 1) to 03 May 2020 (i.e. *n* = 64), from Model-1, using the parameter values obtained by fitting the model to the actual data (Fig. 1). For the case of *no lockdown imposition*, we have taken *a = b =* 0.2. As per prediction of this model, the number (*z_n_*) would have been nearly 10 times larger than its recorded value, on the 64^th^ day, if the lockdown had not been in effect from the 25^th^ day onwards.

**Figure 7:**
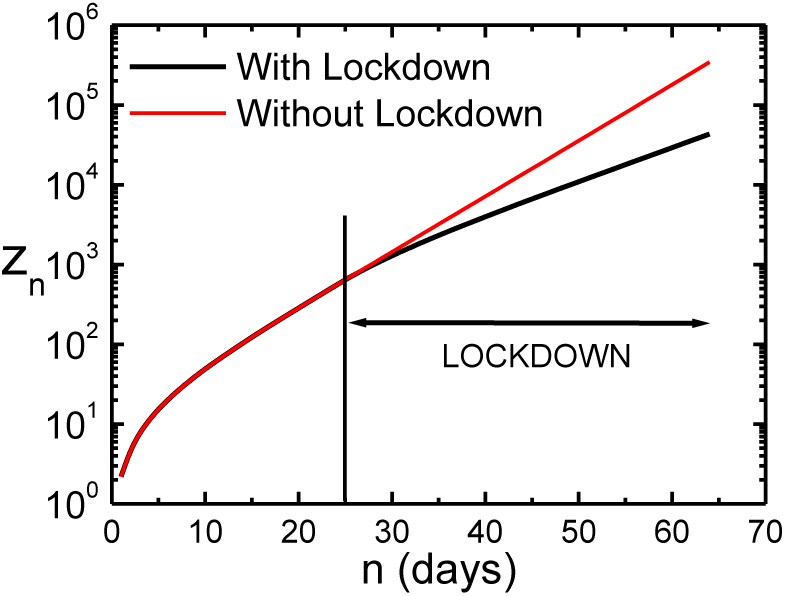
Variation of the number of symptomatic patients as a function of time, under lockdown (black) and without lockdown (red), over the period from 01 March 2020 (i.e. *n* = 1) to 03 May 2020 (i.e. *n* = 64), from Model-1, using the parameter values obtained by fitting the model to the actual data (Fig. 1).

Figure 8 shows the variation of the number of symptomatic patients as a function of time, with and without the imposition of lockdown, over the period from 01 March 2020 (i.e. *n* = 1) to 03 May 2020 (i.e. *n* = 64), from Model-2, using the parameter values obtained by fitting the model to the actual data (Fig. 2). For the case of *no lockdown imposition*, we have taken *a = b =* 0.2. As per prediction of this model, the number (*z_n_*) would have been nearly 10 times larger than its recorded value, on the 64^th^ day, if the lockdown had not been in effect from the 25^th^ day onwards.

Figure 9 shows the variation of the number of symptomatic patients as a function of time, with and without the imposition of lockdown, over the period from 01 March 2020 (i.e. *n* = 1) to 03 May 2020 (i.e. *n* = 64), from Model-3, using the parameter values obtained by fitting the model to the actual data (Fig. 3). For the case of *no lockdown imposition*, we have taken *a = b =* 0.2. As per prediction of this model, the number (*z_n_*) would have been nearly 10 times larger than its recorded value, on the 64^th^ day, if the lockdown had not been in effect from the 25^th^ day onwards.

**Figure 8:**
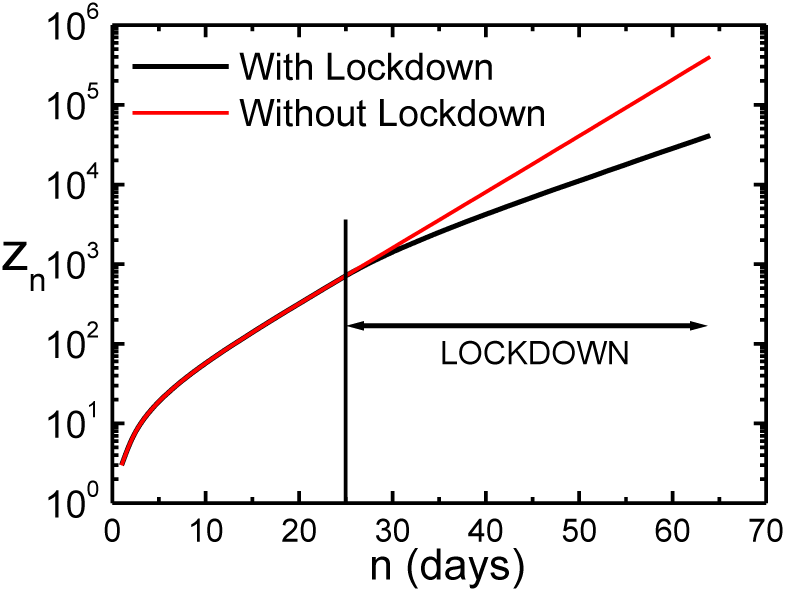
Variation of the number of symptomatic patients as a function of time, under lockdown (black) and without lockdown (red), over the period from 01 March 2020 (i.e. *n* = 1) to 03 May 2020 (i.e. *n* = 64), from Model-2, using the parameter values obtained by fitting the model to the actual data (Fig. 2).

**Figure 9:**
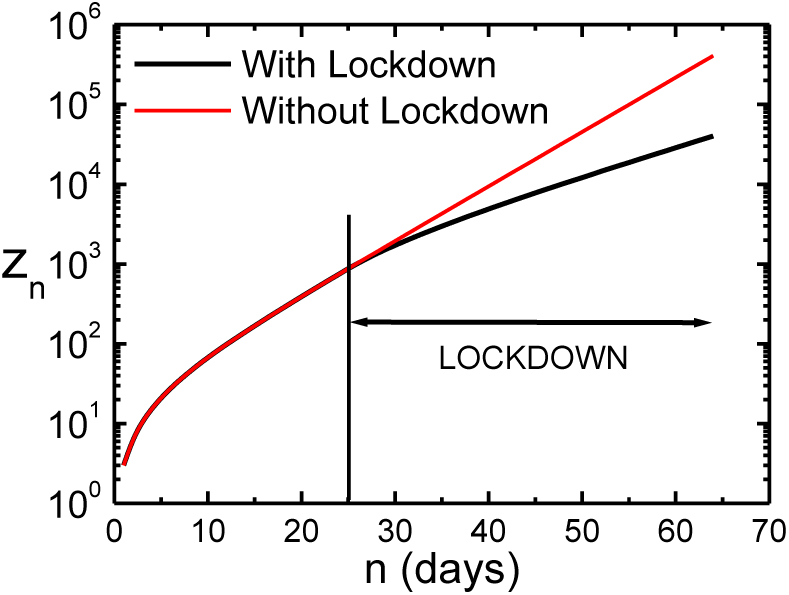
Variation of the number of symptomatic patients as a function of time, under lockdown (black) and without lockdown (red), over the period from 01 March 2020 (i.e. *n* = 1) to 03 May 2020 (i.e. *n* = 64), from Model-3, using the parameter values obtained by fitting the model to the actual data (Fig. 3).

Figure 10 shows the time variation of the number of symptomatic patients, in three phases: 1) pre lockdown, 2) during lockdown, 3) post lockdown, from Model-1, where the lockdown continues till 17 May 2020 (as per the announcement made on 01 may 2020), using the parameter values obtained by fitting the model to the actual data (Fig. 1). It has been assumed here that lockdown won’t continue beyond 17 May 2020. Here *n =* 1 for 01 March 2020.

**Figure 10:**
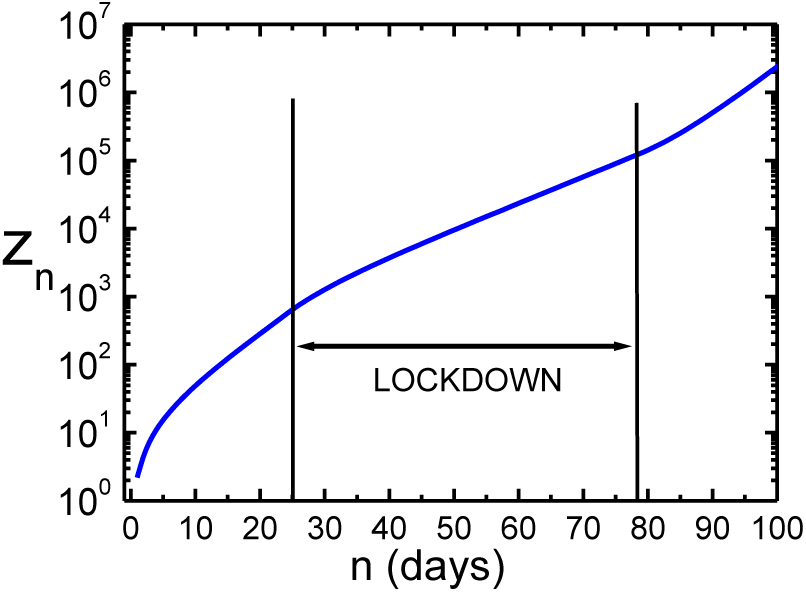
Variation of the number of symptomatic patients as a function of time, for three phases: 1) pre lockdown, 2) during lockdown, 3) post lockdown, from Model-1. The lockdown is to continue till 17 May 2020, i.e. *n* = 78, (as per the announcement made on 01 May 2020). Here we have *n* = 1 for 01 March 2020.

Figure 11 shows the time variation of the number of symptomatic patients, in three phases: 1) pre lockdown, 2) during lockdown, 3) post lockdown, from Model-2, where the lockdown continues till 17 May 2020 (as per the announcement made on 01 may 2020), using the parameter values obtained by fitting the model to the actual data (Fig. 2). It has been assumed here that lockdown won’t continue beyond 17 May 2020. Here *n =* 1 for 01 March 2020.

**Figure 11:**
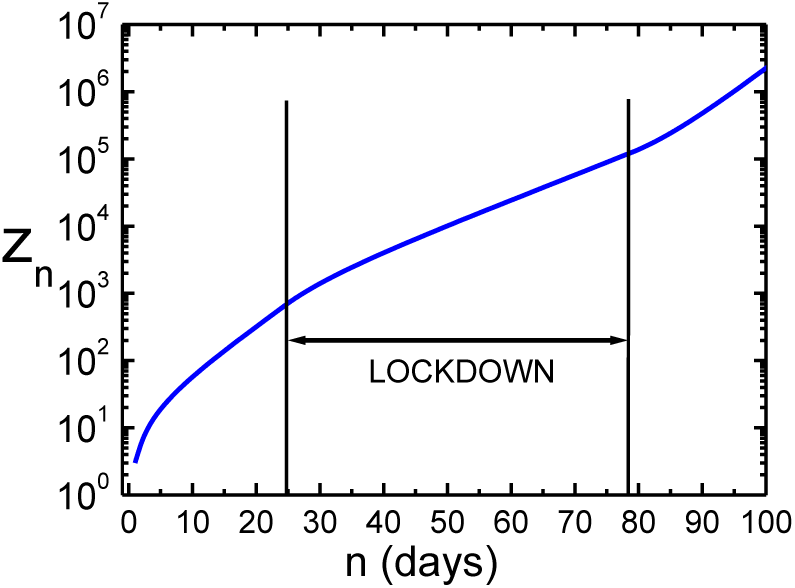
Variation of the number of symptomatic patients as a function of time, for three phases: 1) pre lockdown, 2) during lockdown, 3) post lockdown, from Model-2. The lockdown is to continue till 17 May 2020, i.e. *n* = 78, (as per the announcement made on 01 May 2020). Here we have *n* = 1 for 01 March 2020.

Figure 12 shows the time variation of the number of symptomatic patients, in three phases: 1) pre lockdown, 2) during lockdown, 3) post lockdown, from Model-3, where the lockdown continues till 17 May 2020 (as per the announcement made on 01 may 2020), using the parameter values obtained by fitting the model to the actual data (Fig. 3). It has been assumed here that lockdown won’t continue beyond 17 May 2020. Here *n* = 1 for 01 March 2020.

**Figure 12:**
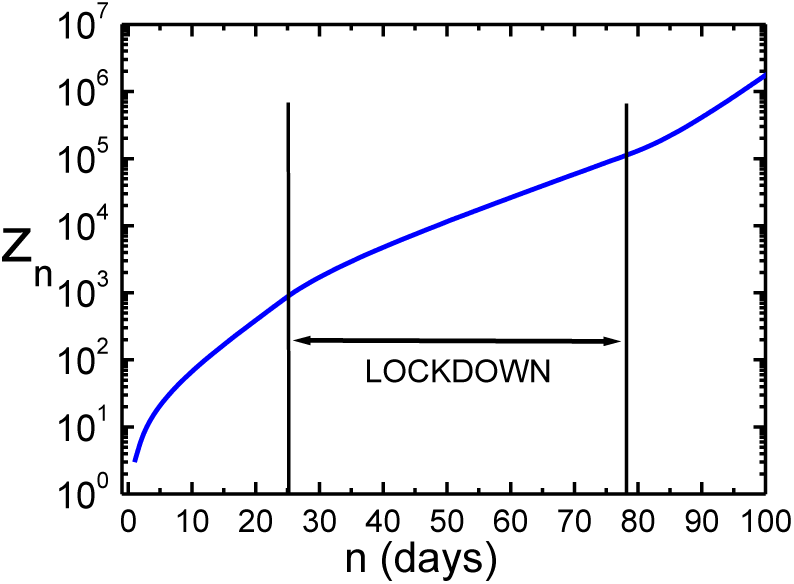
Variation of the number of symptomatic patients as a function of time, for three phases: 1) pre lockdown, 2) during lockdown, 3) post lockdown, from Model-3. The lockdown is to continue till 17 May 2020, i.e. *n* = 78, (as per the announcement made on 01 May 2020). Here we have *n* = 1 for 01 March 2020.

Figure 13 shows the time evolution of the number of asymptomatic patients from Model-1, for three values of the parameter *b*, over a period of 100 days, where lockdown continues from the 25^th^ day onwards. These three values of *b* are smaller than the value obtained by fitting this model to the recorded data (Fig. 1). A smaller value of *b* means greater degree of strictness in enforcing the lockdown. This figure shows that, for a sufficiently small value of *b*, *y _n_* starts decreasing just after the imposition of lockdown.

**Figure 13:**
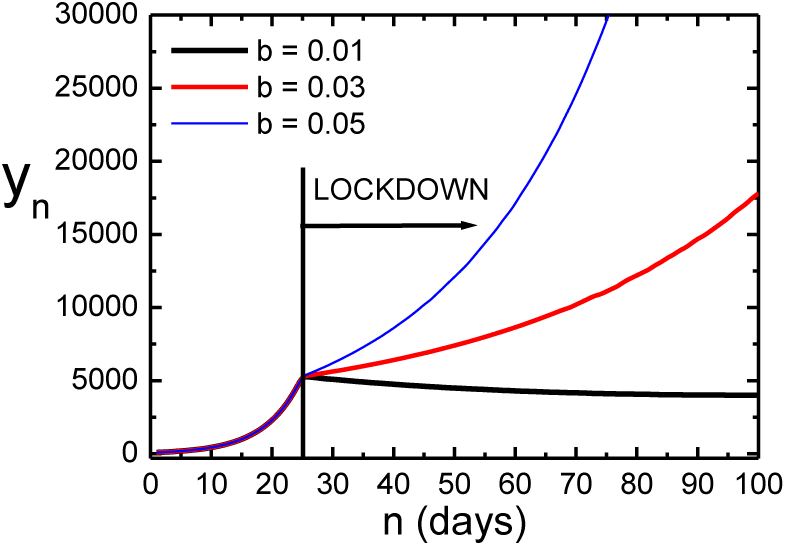
Variation of the number of asymptomatic patients as a function of time for three values of the parameter b, from Model-1, over a period of 100 days, assuming the lockdown to continue from the 25^th^ day onwards. A smaller value of *b* means greater degree of social distancing. Here we have *n* = 1 for 01 March 2020.

Figure 14 shows the time evolution of the number of symptomatic patients from Model-1, for three values of the parameter *b*, over a period of 100 days, where lockdown continues from the 25^th^ day onwards. These three values of *b* are smaller than the value obtained by fitting this model to the recorded data (Fig. 1). A smaller value of *b* means a greater degree of social distancing in the lockdown. This figure shows that, for a sufficiently small value of *b*, the slope of starts decreasing just after the imposition of lockdown, approaching gradually a constant value of *z_n_*. A constant value of *z_n_* means no new cases of symptomatic patients are reported.

**Figure 14:**
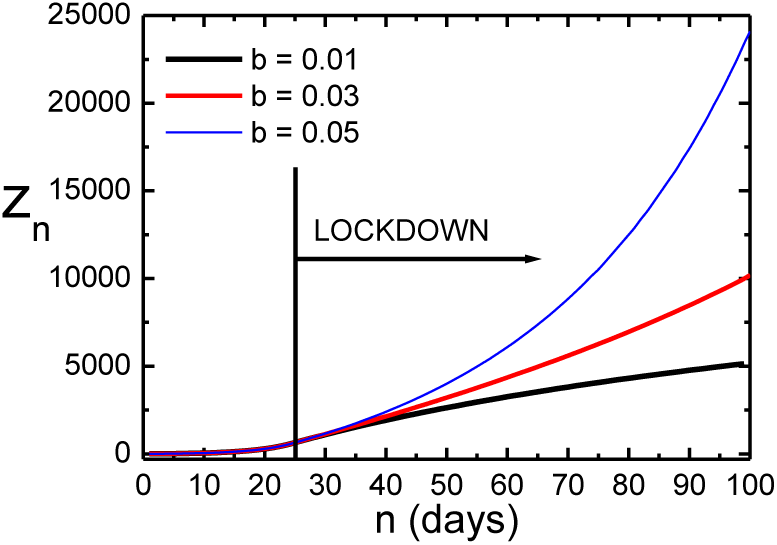
Variation of the number of symptomatic patients as a function of time for three values of the parameter *b*, from Model-1, over a period of 100 days, assuming the lockdown to continue from the 25^th^ day onwards. A smaller value of *b* means greater degree of social distancing. Here we have *n* = 1 for 01 March 2020.

Figure 15 shows the time evolution of the number of asymptomatic patients from Model-2, for three values of the parameter *b*, over a period of 100 days, where lockdown continues from the 25^th^ day onwards. These three values of *b* are smaller than the value obtained by fitting this model to the recorded data (Fig. 2). A smaller value of *b* means a greater degree of social distancing in the lockdown. This figure shows that, for a sufficiently small value of *b*, *y_n_* starts decreasing just after the imposition of lockdown.

**Figure 15:**
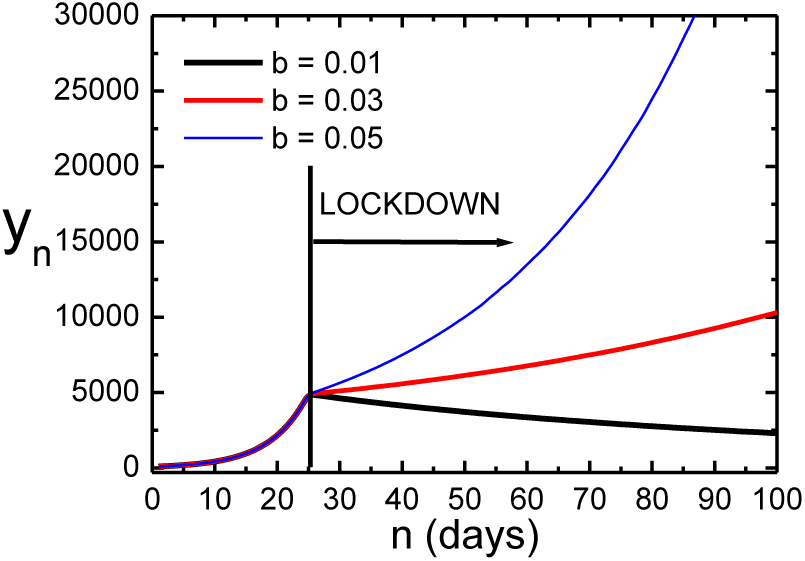
Variation of the number of asymptomatic patients as a function of time for three values of the parameter *b*, from Model-2, over a period of 100 days, assuming the lockdown to continue from the 25^th^ day onwards. A smaller value of *b* means greater degree of social distancing. Here we have *n* = 1 for 01 March 2020.

Figure 16 shows the time evolution of the number of symptomatic patients from Model-2, for three values of the parameter *b*, over a period of 100 days, where lockdown continues from the 25^th^ day onwards. These three values of *b* are smaller than the value obtained by fitting this model to the recorded data (Fig. 2). A smaller value of *b* means greater degree of strictness in enforcing the lockdown. This figure shows that, for a sufficiently small value of *b*, the slope of *z_n_* starts decreasing just after the imposition of lockdown, approaching gradually a constant value of *z_n_*. A constant value of *z_n_* means no new cases of symptomatic patients are reported.

Figure 17 shows the time evolution of the number of asymptomatic patients from Model-3, for three values of the parameter *b*, over a period of 100 days, where lockdown continues from the 25^th^ day onwards. These three values of *b* are smaller than the value obtained by fitting this model to the recorded data (Fig. 3). A smaller value of *b* means a greater degree of social distancing in the lockdown. This figure shows that, for a sufficiently small value of *b*, *y_n_* starts decreasing just after the imposition of lockdown.

**Figure 16:**
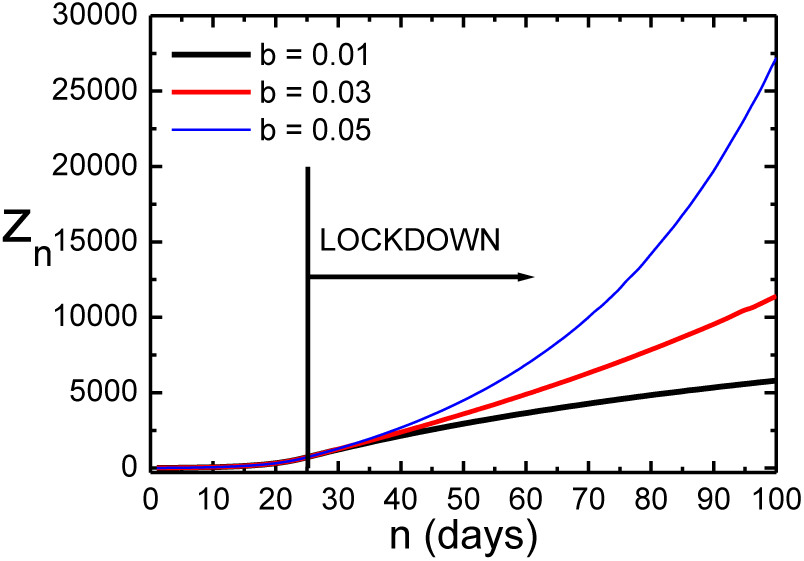
Variation of the number of symptomatic patients as a function of time for three values of the parameter *b*, from Model-2, over a period of 100 days, assuming the lockdown to continue from the 25^th^ day onwards. A smaller value of *b* means greater degree of social distancing. Here we have *n* = 1 for 01 March 2020.

**Figure 17:**
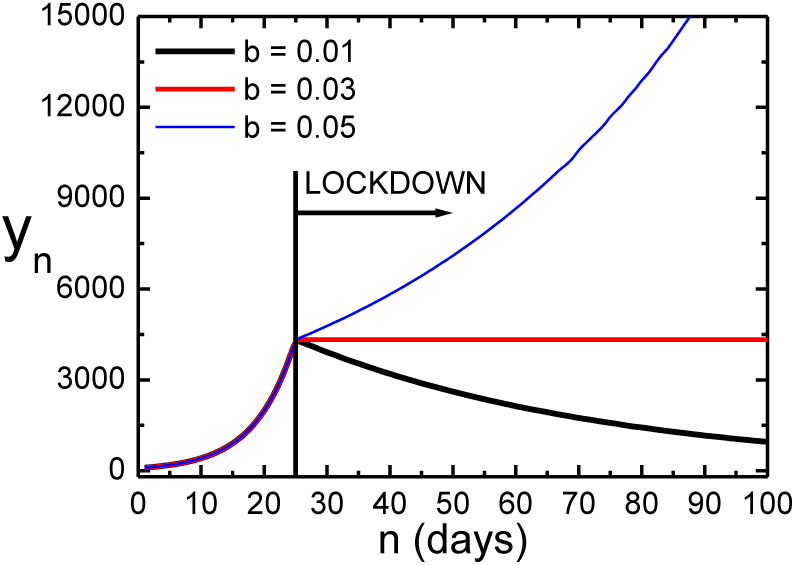
Variation of the number of asymptomatic patients as a function of time for three values of the parameter *b*, from Model-3, over a period of 100 days, assuming the lockdown to continue from the 25^th^ day onwards. A smaller value of *b* means greater degree of social distancing. Here we have *n* = 1 for 01 March 2020.

Figure 18 shows the time evolution of the number of symptomatic patients from Model-3, for three values of the parameter *b*, over a period of 100 days, where lockdown continues from the 25^th^ day onwards. These three values of *b* are smaller than the value obtained by fitting this model to the recorded data (Fig. 3). A smaller value of *b* means greater degree of strictness in enforcing the lockdown. This figure shows that, for a sufficiently small value of *b*, the slope of *z_n_* starts decreasing just after the imposition of lockdown, approaching gradually a constant value of *z_n_*. A constant value of *z_n_* means no new cases of symptomatic patients are reported.

**Figure 18:**
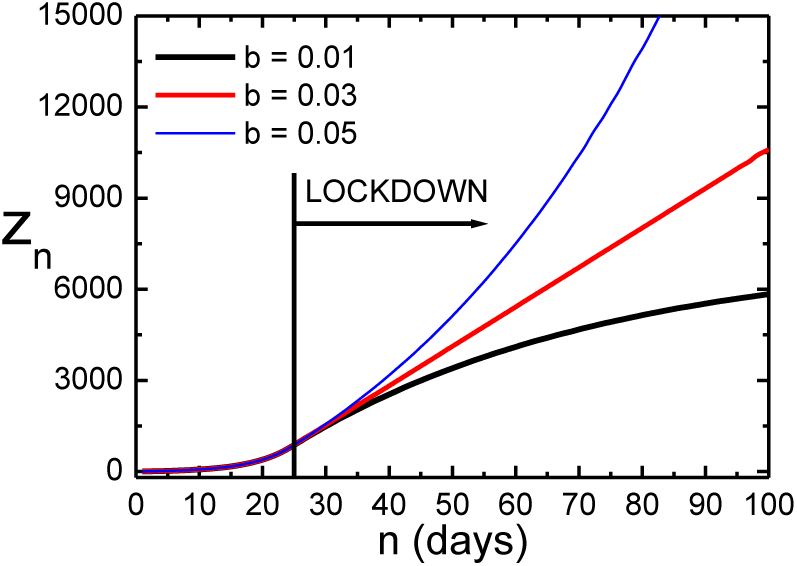
Variation of the number of symptomatic patients as a function of time for three values of the parameter *b*, from Model-3, over a period of 100 days, assuming the lockdown to continue from the 25^th^ day onwards. A smaller value of *b* means greater degree of social distancing. Here we have *n* = 1 for 01 March 2020.

Figure 19 shows the time variation of the relative proportions of asymptomatic (A) and symptomatic (S) patients (expressed as percentage of the total number of patients), based on three models, over a period of 100 days since 01 March 2020 (i.e. *n =* 1), assuming the lockdown to continue till 17 May 2020 (as per the announcement made on 01 May 2020) and not beyond. The smallest value of the ratio of A/S is 3/2. Due to lack of tests in sufficient numbers, the asymptomatic cases mostly remain undetected. Therefore, we may conclude that for every two confirmed cases there are at least three undetected cases in India.

**Figure 19:**
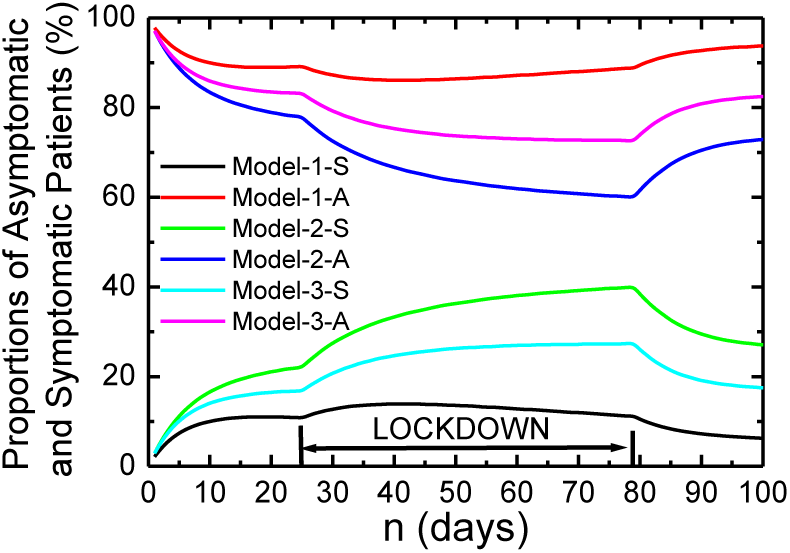
Variation of the relative proportions of asymptomatic (A) and symptomatic (S) patients (expressed as percentages of the total number of patients), as a function of time, based on three models. The lockdown is to continue till 17 May 2020 (as per the announcement made on 01 May 2020) and assumed to be withdrawn thereafter. The smallest value of the ratio of A/S is found to be 3/2. Here we have *n* = 1 for 01 March 2020.

Figure 20 shows the time variation of the percentage of total Indian population infected with COVID-19, with both asymptomatic & symptomatic modes taken together, over a period of 100 days since 01 March 2020 (i.e. *n =* 1), assuming the lockdown to continue till 17 May 2020 (as per the announcement made on 01 May 2020) and not beyond. The present population of India is 1.36 × 10^9^. It shows that on the 78^th^ day (17 May 2020), about 0.1 % of the entire population can be infected, as per the prediction based on Model-1. A smaller percentage is predicted by the other two models.

**Figure 20:**
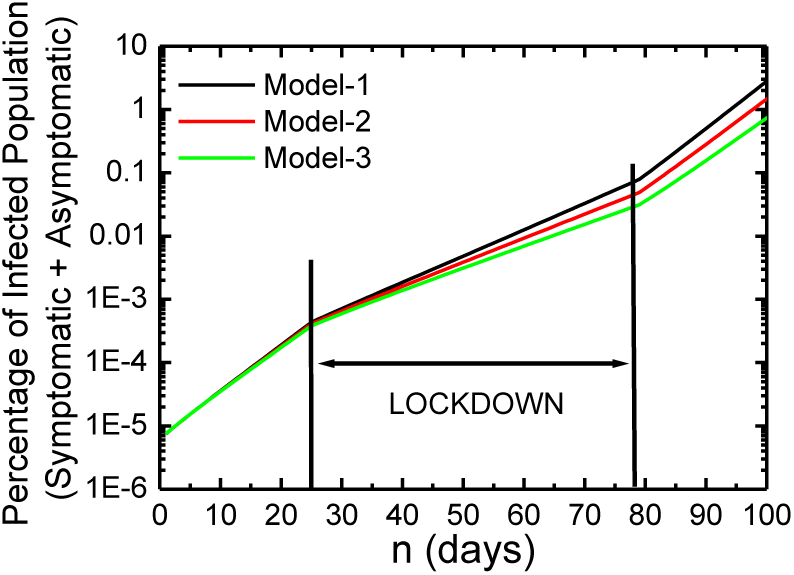
Variation of the percentage of the total Indian population infected, as a function of time, taking asymptomatic & symptomatic cases together, over a period of 100 days since 01 March 2020. The lockdown is to continue till 17 May 2020 (as per the announcement made on 01 May 2020) and assumed to be withdrawn thereafter. The present population of India is 1.36 × 10^9^. Here we have *n* = 1 for 01 March 2020.

## 4. Conclusions

The present study is based on an assumption that a symptomatic patient is put into complete isolation and thereby he/she is prevented completely from spreading the disease. This cannot be entirely true under the present circumstances. The infection in the body of a person, who has developed some symptoms, can remain undiagnosed mainly due to two reasons. One of the reasons is that some of the symptoms are very much similar to those of other diseases (caused by influenza viruses). The other reason is obviously the lack of testing facilities in the country. This method of algebraic study or prediction can be improved by taking into consideration the role played by the symptomatic patients in the transmission of the disease. Another aspect, which has a plenty of scope for modification, is the functional form of *β_j_* of equation (6). Apart from the three choices, in this regard, described in the sections 2.1-2.3, there can be many other functions that can represent the time dependence of this parameter. We have chosen the simplest ones. A limitation of this calculation is that the values of the parameters *a* and *b*, have been assumed to remain constant over the normal (i.e. pre/postlockdown) period and the lockdown period respectively. In reality, the social mixing or distancing patterns may vary frequently with time during a pandemic. In spite of such limitations, the predictions from these models are in reasonable agreement with the actual records, for a certain set of parameter values. Based on this set, the most important finding of the present study is actually a message that social distancing has to be maintained as stringently as possible, which is quite evident from the Figures 13-18. The value of the parameter *b*, which is an indicator of social distancing during lockdown, needs to be sufficiently decreased, to cause *y_n_* to fall with time and also to get a flat curve for *z_n_*.

## Data Availability

For the present study we have used the data regarding the number of confirmed COVID-19 cases in India during the period from 01 March 2020 to 03 May 2020. These data have been obtained from the website of the Ministry of Health and Family Welfare, Government of India. Apart from this, we have used the website of the World Health Organization (WHO) to obtain the present number of COVID-19 infections in the world.
These sources of data have been mentioned in the article and we have provided their web-links in the 3rd and 14th entries of the section of 'References' at the end of the paper.

## References

[1] D. S. Hui et al. The continuing 2019-nCoV epidemic threat of novel coronaviruses to global health -The latest 2019 novel coronavirus outbreak in Wuhan, China. International Journal of Infectious Diseases, 91:264–266, 2020. https://doi.org/10.1016/j.ijid.2020.01.009

[2] J. Yang et al. Prevalence of comorbidities and its effects in patients infected with SARS-CoV-2: a systematic review and meta-analysis. International Journal of Infectious Diseases, 94: 91–95, 2020. https://doi.org/10.1016/j.ijid.2020.03.017

[3] Website of the World Health Organization (WHO). https://www.who.int/

[4] A. Savarino et al. Effects of chloroquine on viral infections: an old drug against today’s diseases? THE LANCET Infectious Diseases, 3(11):722–727, 2003. https://doi.org/10.1016/s1473-3099(03)00806-5

[5] P. Colson et al. Chloroquine for the 2019 novel coronavirus SARS-CoV-2. International Journal of Antimicrobial Agents, 55(3):105923, 1–2, 2020. https://doi.org/10.1016/j.ijantimicag.2020.105923

[6] J. Gu et al. COVID-19: Gastrointestinal Manifestations and Potential Fecal-Oral Transmission, Gastroenterology, 158:1518–1519, 2020. https://doi.org/10.1053/j.gastro.2020.02.054

[7] J. Liu et al. Community Transmission of Severe Acute Respiratory Syndrome Coronavirus 2, Shenzhen, China, 2020. Emerging Infectious Diseases, 26(6), 2020. https://doi.org/10.3201/eid2606.200239

[8] J. F. Chan et al. A familial cluster of pneumonia associated with the 2019 novel coronavirus indicating person-to-person transmission: a study of a family cluster. The Lancet, 395 (10223): 514–523, 2020. https://doi.org/10.1016/S0140-6736(20)30154-9

[9] Q. Li et al. Early Transmission Dynamics in Wuhan, China, of Novel CoronavirusInfected Pneumonia. The New England Journal of Medicine, 382:1199–1207, 2020. https://doi.org/10.1056/NEJMoa2001316

[10] C. Huang et al. Clinical features of patients infected with 2019 novel coronavirus in Wuhan, China. The Lancet, 395(10223):497–506, 2020. https://doi.org/10.1016/S0140-6736(20)30183-5

[11] R. M. Burke et al. Active Monitoring of Persons Exposed to Patients with Confirmed COVID-19 - United States. Morbidity and Mortality Weekly Report, 69(9):245–246, 2020. https://doi.org/10.15585/mmwr.mm6909e1

[12] S. W. X. Ong et al. Air, Surface Environmental, and Personal Protective Equipment Contamination by Severe Acute Respiratory Syndrome Coronavirus 2 (SARS-CoV-2) From a Symptomatic Patient. Journal of the American Medical Association, 323(16): 1610–1612, 2020. https://doi.org/10.1001/jama.2020.3227

[13] Scientific Brief, Published by the *World Health Organization* (WHO). Title: Modes of transmission of virus causing COVID-19: implications for IPC precaution recommendations. Published on 29 March 2020. WHO reference number: WHO/2019-nCoV/Sci_Brief/Transmission_modes/2020.2 Available online at: https://www.who.int/news-room/commentaries/detail/modes-of-transmission-of-virus-causing-covid-19-implications-for-ipc-precaution-recommendations

[14] Website of the Ministry of Health and Family Welfare, Government of India. https://www.mohfw.gov.in/

[15] P Chatterjee et al. The 2019 novel coronavirus disease (COVID-19) pandemic: A review of the current evidence. Indian Journal of Medical Research, 151(2): 147–159, 2020. https://doi.org/10.4103/ijmr.IJMR_519_20

[16] A Agarwal et al. Guidance for building a dedicated health facility to contain the spread of the 2019 novel coronavirus outbreak. Indian Journal of Medical Research, 151(2): 177–183, 2020. https://doi.org/10.4103/ijmr.IJMR_518_20

[17] S Mandal et al. Prudent public health intervention strategies to control the coronavirus disease 2019 transmission in India: A mathematical model-based approach. Indian Journal of Medical Research, 151(2): 190–199, 2020. https://doi.org/10.4103/ijmr.IJMR_504_20

[18] D. Biswas and S. Roy. Analyzing COVID-19 pandemic with a new growth model for population ecology. (A preprint from www.researchgate.net) https://doi.org/10.13140/RG.2.2.34847.92324/1

[19] Arti M. K. and K. Bhatnagar. Modeling and Predictions for COVID 19 Spread in India. (A preprint from www.researchgate.net). https://doi.org/10.13140/RG.2.2.11427.81444

[20] T. Kaur et al. Anticipating the novel coronavirus disease (COVID-19) pandemic. (A preprint from www.medrxiv.org) https://doi.org/10.1101/2020.04.08.20057430

[21] S. Mukhopadhyay and D. Chakraborty. Estimation of undetected COVID-19 infections in India. (A preprint from www.medrxiv.org) https://doi.org/10.1101/2020.04.20.20072892

[22] A. Rajesh et al. COVID-19 prediction for India from the existing data and SIR(D) model study. (A preprint from www.medrxiv.org) https://doi.org/10.1101/2020.05.05.20085902

[23] R. Ranjan. Predictions for COVID-19 Outbreak in India Using Epidemiological Models. (A preprint from www.medrxiv.org) https://doi.org/10.1101/2020.04.02.20051466

[24] S. Ghosh. Predictive Model with Analysis of the Initial Spread of COVID-19 in India. (A preprint from www.medrxiv.org) https://doi.org/10.1101/2020.05.02.20088997

[25] M. Bhatnagar. COVID-19: Mathematical Modeling and Predictions. (A preprint from www.researchgate.net). https://doi.org/10.13140/RG.2.2.29541.96488

[26] S. Roy and K. Roy Bhattacharya. Spread of COVID-19 in India: A Mathematical Model. Journal of Science and Technology, 5(3): 41–47, 2000. https://doi.org/10.46243/jst.2020.v5.i3.pp41-47

